# A large language model for risk-of-bias assessment in systematic reviews of prognosis studies in clinical neurology

**DOI:** 10.64898/2026.09.09.26362643

**Authors:** Sem L Kampman, Kees PJ Braun, Willem M Otte

**Affiliations:** Department of Child Neurology, UMC Utrecht Brain Center, University Medical Center Utrecht and Utrecht University, Utrecht, The Netherlands, member of ERN EpiCARE

**Keywords:** risk of bias, systematic reviews, prognosis, large language model, neurology

## Abstract

**Background:** Risk-of-bias (ROB) assessments represent an integral component of systematic reviews. However, this task is often highly repetitive, time-consuming, and may lack inter-rater consistency. Large language models (LLMs) offer opportunities for automation in systematic reviews, which may expedite and enhance the quality and consistency of research synthesis.

**Methods:** Using zero-shot prompting, we designed an LLM-based pipeline as a virtual mimic of a human reviewer for the Quality in Prognosis Studies (QUIPS) framework. Then, focusing on prognostic research in a single discipline (neurology), we applied this pipeline to articles included in previously published systematic reviews. We studied inter-rater agreement between both (1) the LLM and the original human ROB assessments and (2) between original human ROB assessments.

**Results:** 298 articles from 15 reviews across three domains (epilepsy, traumatic brain injury, stroke) were included. We demonstrate the feasibility of a tailored, prompt-engineered LLM pipeline for automating ROB assessments with the QUIPS tool. While LLM-human agreement was limited (Cohen’s weighted kappa; = 0.22, 95% CI, 0.12 - 0.33), our data tentatively suggest, based on a small sample (n=5), that it may not be inferior to human-human agreement (Cohen’s weighted kappa; = -0.25, 95% CI, -1.04 - 0.54). Wilcoxon signed-rank tests were statistically significant (p < 0.05) across four bias domains and for the overall risk scores, and rank-biserial correlations demonstrated human raters’ tendency to assign higher risk scores than LLM counterparts.

**Conclusions:** With targeted methodological refinements - including standardization of QUIPS implementation and validation against expert ratings - automated ROB assessments may meaningfully reduce time and cost of systematic reviews of prognosis studies in neurology and beyond.

## Introduction

Systematic reviews and meta-analyses occupy the highest tier of evidence synthesis in medical science ^2,3^. Because findings commonly inform guidelines ^2,4^, rigorous standards are essential to assure methodologically sound reviews ^5^. Systematic reviews also contribute to the development of clinical tools ^6^. Candidate predictor variables identified in reviews may inform clinical prediction models, such as for estimating seizure recurrence ^6–9^. In neurology and beyond, prognostic research is a priority area of focus, given its central role in shared decision-making, biomarker discovery, and personalized medicine ^10–12^. Moreover, as common neurological conditions grow more prevalent, such as Parkinson’s disease and stroke ^13,14^, the global burden of neurological disorders continues to rise ^15^, underscoring the importance of high-quality evidence synthesis in this field ^16^.

A key component of systematic reviews is risk-of-bias (ROB) assessment. This step assesses internal validity and bias in included studies ^17^, which may help explain between-study heterogeneity and serve to qualify study conclusions ^18^. Articles deemed highly biased may downgrade the overall reliability of findings or may be removed from the primary analysis or systematic review altogether ^17,19–21^.

ROB assessment remains a tedious and time-consuming endeavor, with formal recommendations specifying two independent reviewers ^22^. Furthermore, despite requiring substantial methodological expertise for consistent scoring ^23^, this task may not always receive the same level of scrutiny as other methodological components. As a result, ROB scores may lack accuracy and suffer from internal and external inconsistency ^24–27^.

Ongoing advancements in machine learning have enabled (semi-)automated approaches for systematic reviews ^5,28,29^. Tools exist for streamlining article screening and selection ^30,31^ and automating data extraction ^29,32^. Besides conserving time and resources, automated ROB tools have been developed in efforts to improve the quality and consistency of assessments ^28,33–35^. More recently, large-language models (LLMs) have been studied ^1,36–39^, especially in systematic reviews of randomized clinical trials (e.g., using the ROB-2 tool) ^39–42^. Benefits of LLMs include their user friendliness, adaptability, and ability to analyze voluminous datasets^36,43^, although concerns remain regarding the tendency to “hallucinate” ^44^.

To our knowledge no prior effort has automated ROB assessment using LLMs with the Quality in Prognosis Studies (QUIPS) tool ^45^, or in neurology systematic reviews specifically. While the potential utility of integrating artificial intelligence in clinical neurology care receives growing interest ^46,47^, its role in evidence synthesis has garnered comparatively little attention.

### Study Aims

Here, we designed an LLM-based ROB-assessment pipeline using the QUIPS tool for systematic reviews of prognosis studies in three common neurological conditions: epilepsy, traumatic brain injury (TBI) and stroke. First, we analyzed inter-rater agreement and group-level differences between published human assessments and LLM assessments. Then, we calculated agreement between original human-human ratings – for studies published in more than one included systematic review – to explore the “gold-standard” or human reference standard of agreement.

## Methods

Ethical approval was not required as we only analyzed published meta-data. Where applicable, we followed journal guidance on reporting for studies evaluating the use of generative A.I. in research synthesis ^48^. However, because the field of LLM-assisted ROB assessment is in its infancy, specific reporting guidelines are absent - this represents a crucial area of development ^1,36^.

### Systematic review selection

PubMed was searched (February 2026) to identify published prognostic systematic reviews that used the QUIPS Tool, in three common areas of clinical neurology: epilepsy, traumatic brain injury (TBI) and stroke (see **S1A**). We selected an exploratory, convenience sample of 15 systematic reviews — five per neurological domain (epilepsy, TBI, stroke) — published in the last ∼15 years. Inclusion required application of the QUIPS tool and accessibility of original article-level ratings; we did not perform a comprehensive meta-review. Search terms combined the tool name (QUIPS) with domain keywords (epilepsy, traumatic brain injury, stroke) and review-type terms (systematic review, prognostic).

### The QUIPS tool

The QUIPS tool provides a framework for evaluating presence of bias across six domains in prognostic studies: Study Participation, Study Attrition, Prognostic Factor Measurement, Outcome Measurement, Study Confounding and Statistical Analysis and Reporting ^45,49^; for elaboration on domain-specific criteria, as well as the development, underlying rationale and recommended usage of the tool, see ^45,49,50^. Per domain, guided by a set of structured prompting questions, risk can be graded as ‘Low’, ‘Moderate’ or ‘High’ ^49^.

The original QUIPS documentation did not specify how to aggregate risk scores across bias domains for an overall study score, though it advised against “summated scores” ^49^.

Because an overall ROB score is often desired for reporting in systematic reviews, a more recent method was proposed for deriving an overall rating from its constituent domain scores^50,51^:

1. “Low” risk: “Low” ROB across all domains, or maximum one domain with “Moderate” risk

2. “High” risk: “Moderate” ROB across 3 or more domains, or any domain with “High” risk

3. “Moderate” risk: any score that does not meet overall criteria for “Low” or “High” risk

In this work, we maintain this scoring system for aggregation of bias outcomes, overriding existing original aggregate scores when they deviate from our scoring system. For articles not assessing certain domains (e.g., Giuliano et al. ^52^ and West et al. ^53^, see S1), we did not aggregate overall scores to ensure fair comparisons of ratings between LLM and human assessors.

### LLM-Based Assessment Pipeline

#### Step 1: Prompt engineering

We developed a structured system prompt instructing the LLM to assume the role of a “Virtual Clinical Expert and Methodologist specializing in Prognosis Studies.” We adopted this persona-based framing because role assignment is a commonly used prompt-engineering technique in clinical LLM applications ^54^ though evidence for its consistent benefit is mixed ^55^. The dual emphasis on clinical expertise and methodological specialization was intended to balance *content knowledge* (understanding prognostic study designs) with *process knowledge* (applying quality assessment criteria). The prompt embedded explicit methodological principles derived from the QUIPS framework (https://methods.cochrane.org/sites/methods.cochrane.org.prognosis/files/uploads/QUIPS%20tool.pdf), including guidance on evaluating selection bias (sample representativeness, participation rates ≥80% as low risk), study attrition (informative censoring, systematic differences between completers and non-completers), measurement bias (validity, reliability, and assessor blinding), confounding (identification and adjustment for key confounders), and reporting bias (detection of selective reporting and data-dependent variable selection).

To prevent hallucinations, we incorporated a critical distinction between "No" and "Unsure" responses. This distinction addresses a fundamental challenge in ROB assessment: distinguishing between what a study failed to do versus what a study failed to report. “No” was defined as appropriate when a manuscript demonstrably lacked a required element (e.g., no description of the source population), representing a reporting deficit that should negatively influence the risk assessment. “Unsure” was reserved for situations where the manuscript provided insufficient information to form a methodological judgement, representing epistemic uncertainty that should not be conflated with confirmed deficiency. This distinction was implemented to prevent the LLM from penalizing studies for ambiguous reporting when methodological quality might nonetheless be adequate, while still appropriately flagging inadequate reporting transparency.

We instructed the LLM to provide three elements for each assessment item: a rating ("Yes", "Partial", "No", or "Unsure"), a verbatim evidence snippet quoted directly from the manuscript text (or "Not found in text" if absent),^56^ and reasoning explaining how the evidence supported the assigned rating. The complete prompt is available in the supplementary materials and online repository.

#### Step 2: PDF to Markdown conversion

We manually retrieved PDF versions of included articles from the previously published systematic reviews. Prior to assessment, we converted PDFs to Markdown format using marker-single software (marker-pdf package, version 1.0) to enable automated text processing. Conversion parameters disabled image extraction and enabled automatic language detection to accommodate non-English publications. The conversion preserved full manuscript text including methods, results, and discussion sections, along with table structures and figure captions.

#### Step 3: Automated ROB assessment

Markdown files were processed using a custom Python script interfacing with the LLM model through the OpenRouter API gateway. The complete QUIPS schema comprising six domains and 32 items was encoded in JSON format. Note that we had one more prompting item than the original documentation, because we encoded separate prompting items for assessing the description of period and place of recruitment. For each manuscript, the system prompt and full Markdown content were submitted to the LLM, which returned a structured JSON response containing item-level ratings, evidence snippets, reasoning, and synthesized domain-level risk assessments.

#### Step 4: Output and data extraction

We saved LLM outputs as individual JSON files to preserve complete item-level data. A post-processing script converted these to tabulated format (TSV) for statistical analysis. Overall, ROB scores were calculated deterministically during post-processing using the Grooten et al. criteria ^50^, ensuring algorithmic consistency across all assessments.

#### Model selection and configuration

We used Google Gemini-3-Pro-Preview accessed via the OpenRouter API (base URL: https://openrouter.ai/api/v1). Temperature was set to 0·0 to ensure deterministic, reproducible outputs. The response format was constrained to JSON objects to enforce structured output. A rate limit of one request per second was implemented to prevent API throttling. Processing metadata including timestamps, model identifier, and source file paths were appended to each output for audit purposes.

#### Reporting

The pipeline code, system prompts, QUIPS schema, and example outputs are publicly available at https://github.com/SemKampman/QUIPS_Automated_ROB_Assessment.

#### Comparison with manual assessment

Original manual assessments were retrieved from published articles and compiled in an MS Excel form. Importantly, to ensure consistency with the LLM scoring process, we manually applied the Grooten et al. ^50^ criteria for aggregate risk scoring, overriding original overall scores from several of the systematic reviews.

We ensured LLM-derived ratings were consistent with the pre-defined aggregation algorithm ^41,50^. Two reviewers (SK and WO) then manually checked all articles originating from Cochrane systematic reviews with “two-level” differences for overall ROB ^41^ – that is, when one rater assigned a “High” risk when another assigned a “Low” risk - and attempted to resolve these differences, in an unblinded fashion (**S2**). Verification of LLM ratings of “two-level” incongruencies was only feasible for Cochrane reviews because other published human ratings were not accompanied by (adequately informative) supporting statements.

In the main analysis, we used weighted Cohen’s κ coefficients to compute inter-rater agreement between LLM and original human ROB assessments; articles present in more than one review were treated as independent for this step. Cohen’s κ computes inter-rater agreement between pairs of raters for categorical variables, correcting for chance agreement ^57,58^. Weighted Cohen’s κ was chosen to respect the ordinal structure of answer choices, penalizing greater differences in ranks (i.e., between ‘High’ and ‘Low’) in a quadratic manner (“squared weighing”) ^58,59^. This was chosen as “two-level” discrepancies ^41^ - such as incorrectly classifying a high-risk article as a low-risk article - are particularly concerning.

Κappa agreement was interpreted as follows: ≤ 0.00 = “poor agreement”, 0.01 – 0.20 = “slight agreement”, 0.21–0.40 = “fair agreement”, 0.41–0.60 = “moderate agreement”, 0.61–0.80 = “substantial agreement”, and 0.81–0.99 “near perfect agreement” ^59,60^.

In the secondary analysis, Wilcoxon signed-rank tests were performed to test whether the distribution of categorical outcomes differed between the two raters (LLMs vs humans) for the same articles (paired samples). While Cohen’s κ assesses inter-rater agreement on a per-item basis, the Wilcoxon signed-rank test assesses group-level differences in rankings. Additionally, rank-biserial correlation scores were computed ^61^, to estimate the direction of group-level differences. In other words, whereas Wilcoxon signed-rank tests tell you that a difference exists between two groups, the rank-biserial correlation score tells you in *which* direction this difference exist – that is, whether human or LLM ratings are systematically higher or lower.

In a secondary analysis, to evaluate the “gold-standard” human-human inter-rater agreement, we computed Cohen’s κ ratings for articles found in more than one of the included systematic reviews. In fact, the possibility to study this was an important consideration for including multiple reviews on similar topics (from independent research groups) per neurological discipline. Weighted Cohen’s κ coefficients were compared, for overall ROB and across domains.

Statistical significance was evaluated at α = 0.05; given the exploratory nature of the analysis we did not formally adjust for multiple comparisons across the seven Wilcoxon tests, but we report p-values for transparency. All statistical analyses were performed in RStudio using R version 4.5.1 ^62^. Important packages used for analysis include *psych* ^63^ and *rstatix* ^64^ .

## Results

### Study Characteristics

Five previously published systematic reviews on neurological prognostic outcomes were identified per neurological disorder (epilepsy, TBI, stroke), for a total of 15 systematic reviews (**S1A**) ^52,53,65–77^. 298 articles were available (94 epilepsy articles, 79 stroke articles, and 125 TBI articles) for analysis. Study characteristics, and details regarding QUIPS assessment and modifications are summarized in **S1A/B.** 11/15 studies explicitly reported that ROB assessments were done by two reviewers, whereas three studies did not specify, and in one study a second reviewer randomly sampling only 10%. Out of these 11 articles, none reported kappa scores between their two reviewers or initial incongruences requiring resolution by consensus or adjudication by a third reviewer.

### Discrepancies in ROB assessments

Available human to LLM comparisons ranged per domain from 251 to 298, and 245 pairs were available for comparison of the overall risk score. Visual inspection suggested that human raters tended to systematically assign higher (more conservative) ROB scores than the LLM (***Figure 2).*** Two-level step-wise differences between human and LLM ratings were most prevalent in Domains 2 and 6, with humans often scoring higher (***Figure 2***; ***Table* 1**).

**Figure 1.**
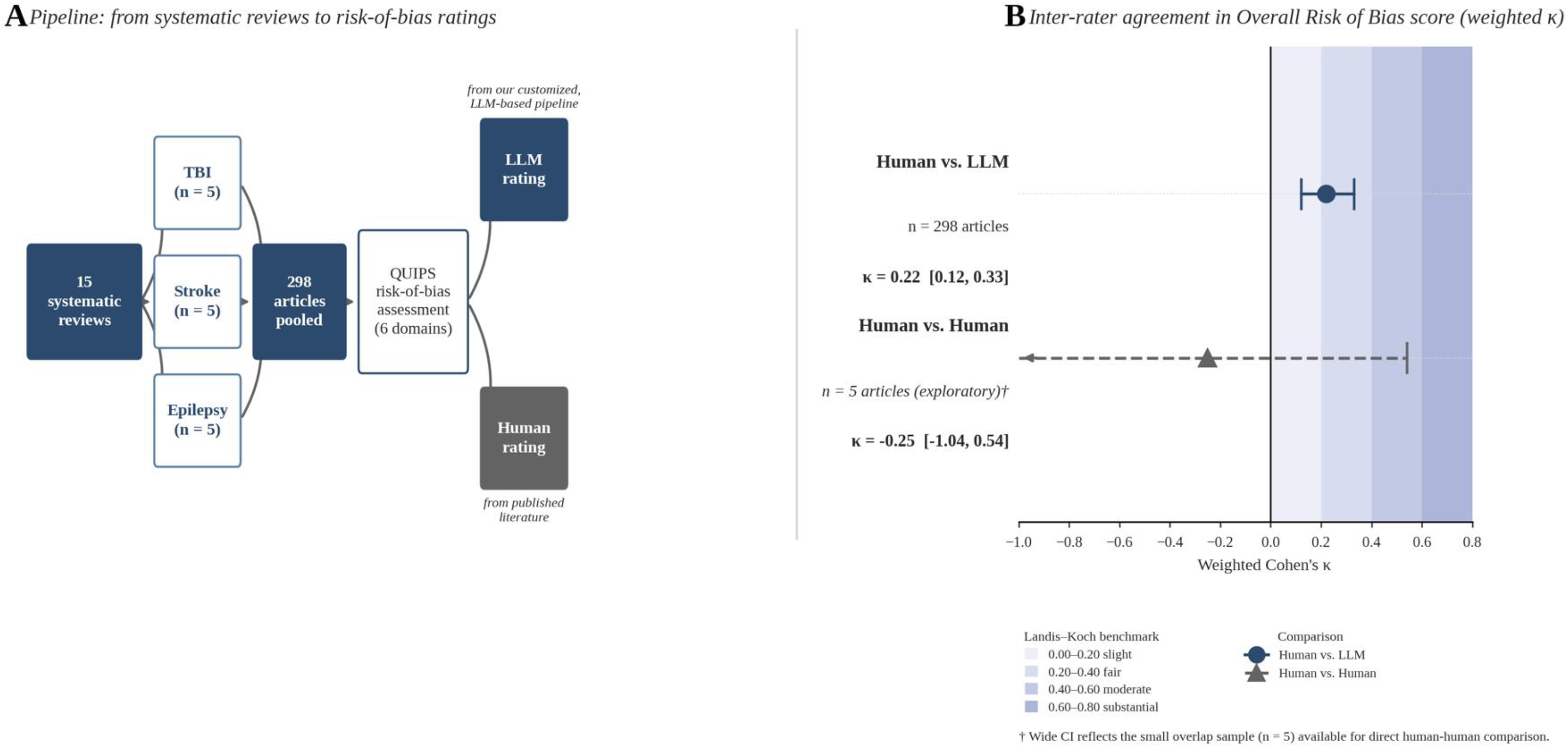
(Graphical Abstract). (A) 298 articles from 15 systematic reviews of neurological conditions were analyzed. Original human QUIPS risk-of-bias ratings were extracted and compared to ratings generated by our customized LLM-based pipeline. (B) Agreement between human and LLM ratings for overall risk-of-bias score between humans and LLMs was limited, although significantly greater than chance, and apparently higher than the human-human agreement calculated from articles included across different systematic reviews. However, caution is urged in interpretations given the extremely limited sample size available (n=5). This figure was created using Claude Sonnet 5 (*Anthropic*).

**Figure 2AB.**
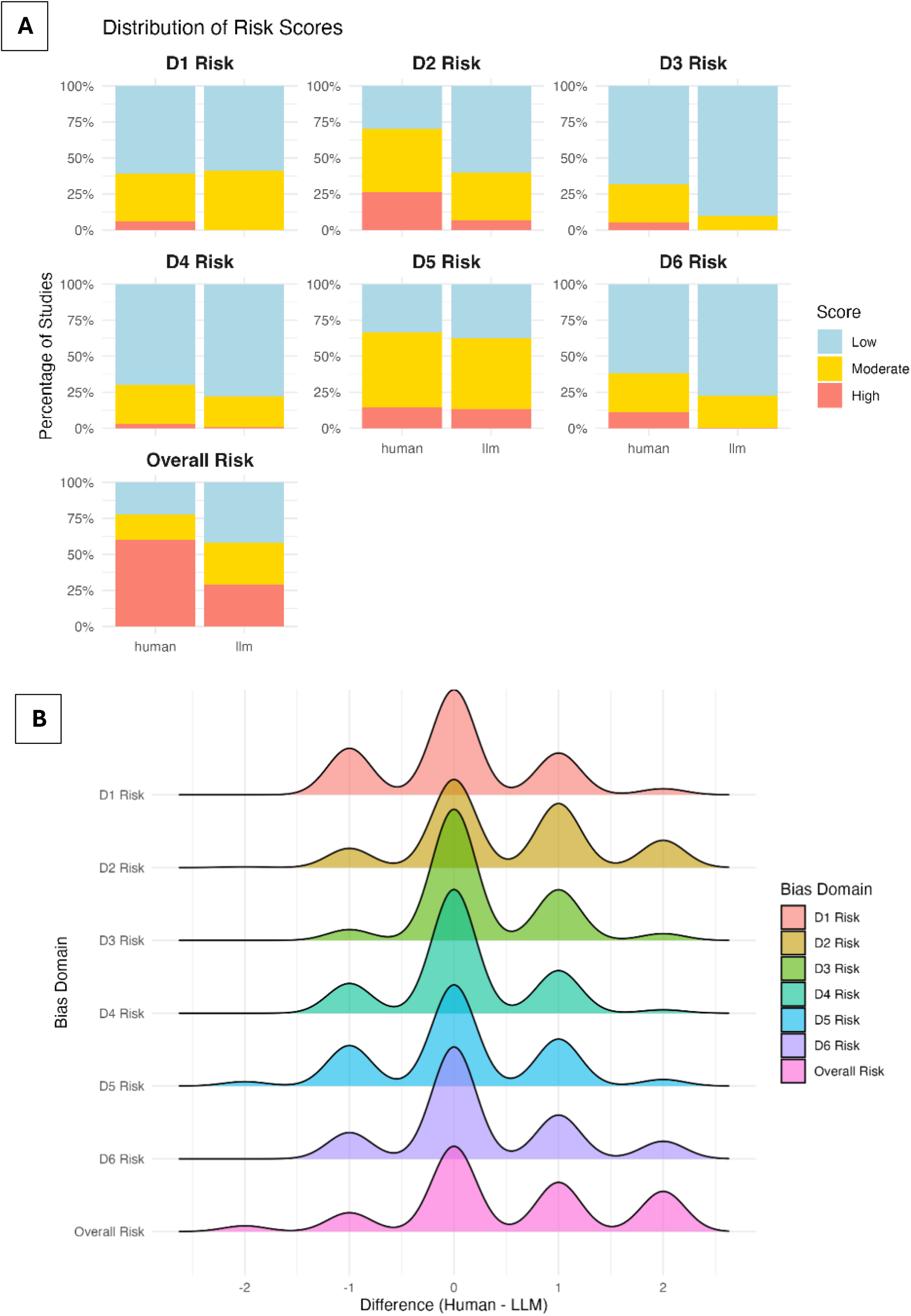
Humans systematically assigned higher ROB scores across domains. Panel A shows raw distributions. Panel B plots how ratings differed between humans and the LLM; for example, a difference of +2 would be present if humans rated a domain as “High” when the LLM rated it as “Low”, and vice versa for a difference of -2.

**Table 1.** “Two-level” mismatches (i.e., differences between “High” and “Low” risk) between human and LLM ratings were most prevalent in Domain 2 and overall risk.

| Bias Domain | Article pairs | # of two-level mismatches | % of two-level mismatches |
| --- | --- | --- | --- |
| D1 Risk | 298 | 9 | 3.0% |
| D2 Risk | 251 | 36 | 14.3% |
| D3 Risk | 297 | 10 | 3.4% |
| D4 Risk | 297 | 5 | 1.7% |
| D5 Risk | 278 | 15 | 5.4% |
| D6 Risk | 298 | 26 | 8.7% |
| **Overall Risk** | 245 | 56 | 22.9% |

Wilcoxon signed-rank test scores were statistically significant across four domains (Domains 2, 3, 4 and 6), as well as for the overall risk score, indicating group-level differences (**Table 2**). The rank-biserial correlations indicate that human raters consistently assigned higher (i.e., more conservative) ROB scores across every domain and for the overall score.

**Table 2.** Overview of Wilcoxon signed-rank tests and rank-biserial correlations across bias domains. Rank biserial correlation scores show lower and upper bounds of 95% confidence intervals. Higher rank-biserial correlation scores (i.e., > 0) indicate systematically higher ratings assigned by human reviewers.

| Bias Domain | Articles | p-value | Rank-biserial correlation (95% C.I.) |
| --- | --- | --- | --- |
| D1 Risk | 298 | 0.353 | 0.017 (0.001–0.140) |
| D2 Risk | 251 | < 0.001 | 0.431 (0.320–0.530) |
| D3 Risk | 297 | < 0.001 | 0.391 (0.300–0.470) |
| D4 Risk | 297 | < 0.01 | 0.141 (0.030–0.250) |
| D5 Risk | 278 | 0.283 | 0.053 (0.002–0.180) |
| D6 Risk | 298 | < 0.001 | 0.295 (0.180–0.390) |
| **Overall Risk** | 245 | < 0.001 | 0.403 (0.300–0.510) |

#### “Two-level” mismatches

We inspected and attempted to resolve “two-level” discrepancies between human and LLM ratings for overall ROB, in an unblinded fashion, for all Cochrane systematic reviews (see **S2**) ^41^. Although we variably agreed with human, LLM or both human and LLM assessments across domains, our overall judgement accorded with the LLM in three of the four examined cases. Generally, the supporting judgements from the LLM were more informative and no evident fabrications were detected; however, given that only 4/56 articles (7%) with two-step differences provided supporting statements enabling inspection and resolution of discrepancies, we cannot extrapolate these findings to all articles.

### Main analysis; weighted Cohen’s κ

Weighted κ between published human and LLM ratings across bias domains ranged from 0.04 to 0.27. Agreement was “slight” in five domains (Domains 1, 2, 3, 4, 6) and “fair” in one domain (Domain 5) (***Table 3***). Lowest agreement was seen in Domain 6. For overall ROB, weighted κ was 0.22 (95% C.I, 0.12-0.33) indicating ‘fair agreement’ per the Landis-Koch scale.

**Table 3.**
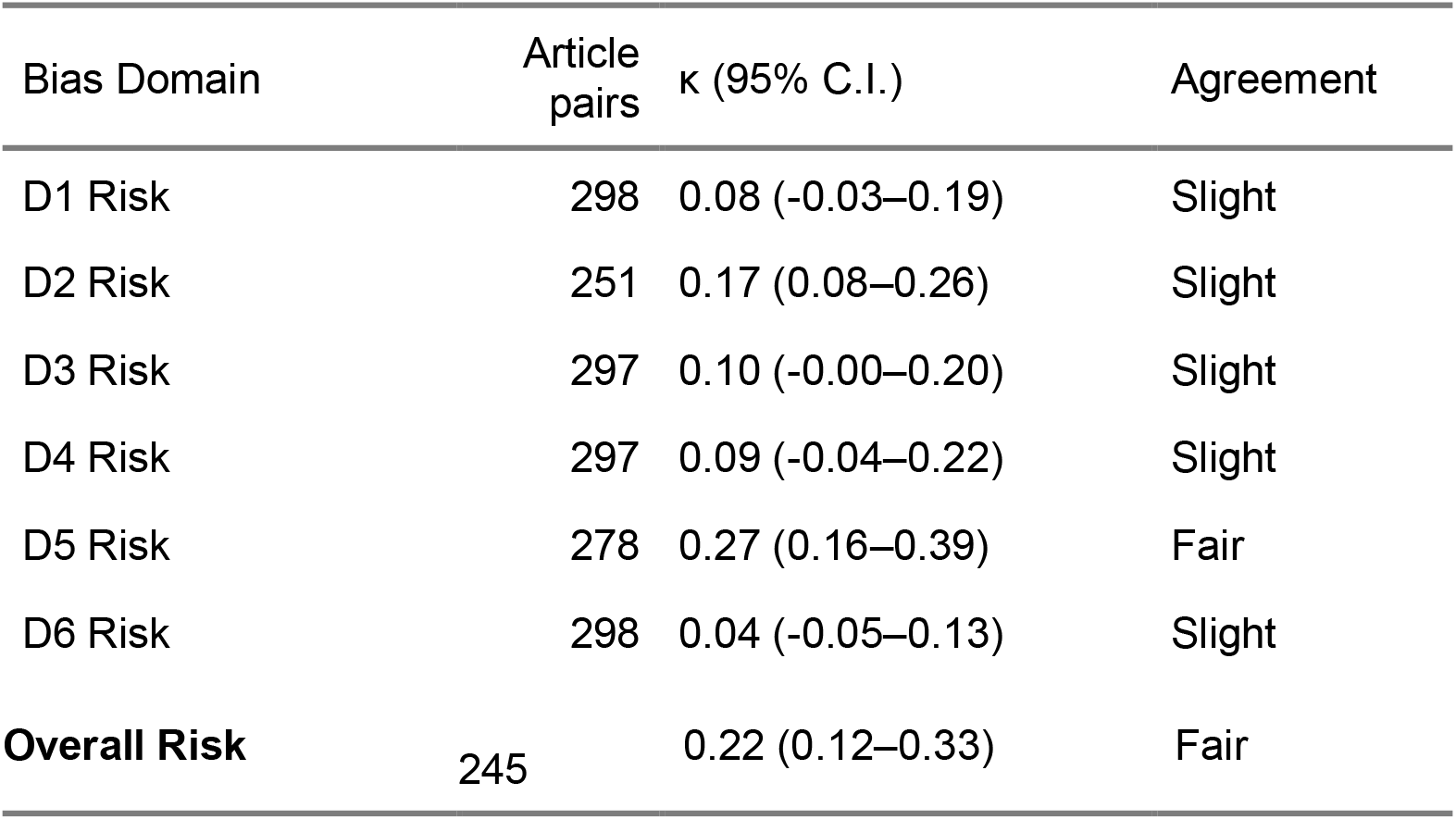
Overview of weighted Cohen’s κ scores across bias domains, penalizing discrepancies between ratings in a quadratic manner. Lowest and highest agreement were seen in Domain 6 and Domain 5, respectively.

Weighted κ for overall risk score between published human and LLM ratings across ranged from -0.15 to 0.44 across systematic reviews. Agreement was poor in six reviews, slight in three reviews, fair in three reviews, and moderate in two reviews (***Table 4***). Two systematic reviews - Giuliano 2021 and West 2019 - consistently rated Domain 2 as “N/A” for retrospective/registry studies, precluding overall risk score aggregation; this explains their absence or underrepresentation in ***Table 4***.

**Table 4.**
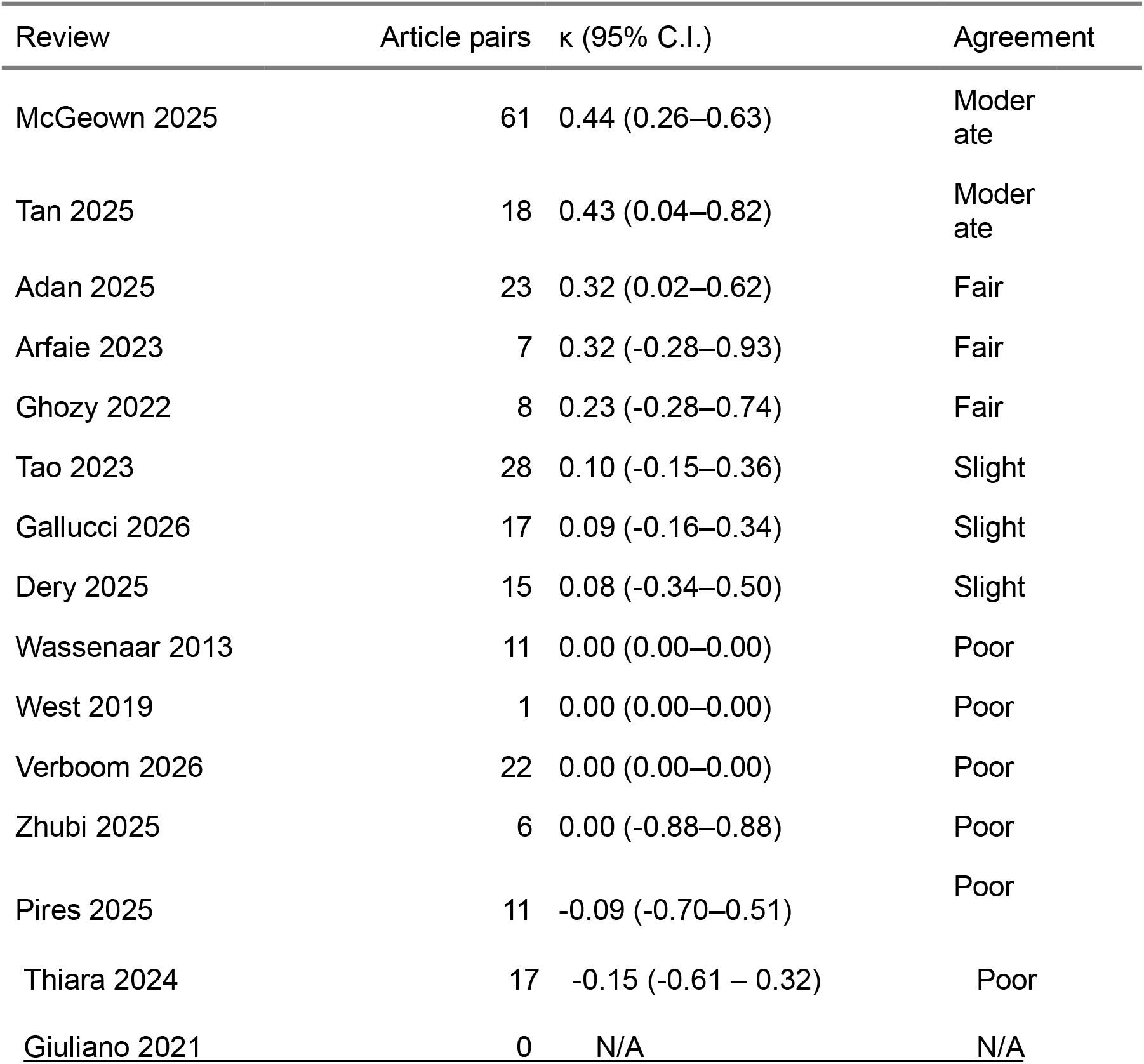
Overview of weighted Cohen’s κ for overall risk scores – i.e., the aggregated ROB ratings across domain using our scoring algorithm-per systematic review, penalizing discrepancies between ratings in a quadratic manner. Overall, agreement between human and LLM ratings was highly variable, ranging from -0.15 to 0.44. **Note:** West et al., and Giuliano et al., did not rate all domains, and hence no aggregated overall risk scores were possible for most or all studies, respectively. For Wassenaar et al., and Verboom et al., overall human risk scores were almost invariably rated as “High”, leaving little variation in the data and therefore producing small agreement estimates and resulting confidence intervals. N/A: not available.

### Human-to-human QUIPS assessments

We found five duplicate articles in our study, enabling analysis of human-to-human agreement. The weighted κ coefficient for human-human pairs (5 articles) for overall risk score was κ -0.25 (95% CI, -1.04–0.54). Thus, while we could conclude that human-LLM agreement for overall ROB was statistically better than chance (κ 0.22, 95% CI, 0.12–0.33), the analysis for the human-human pairs was limited by small sample size (***Table 5***). Finally, for all five of these duplicate articles, the LLM assigned the exact same ratings across all six domains and for overall risk, demonstrating internal consistency (data not shown).

**Table 5.**
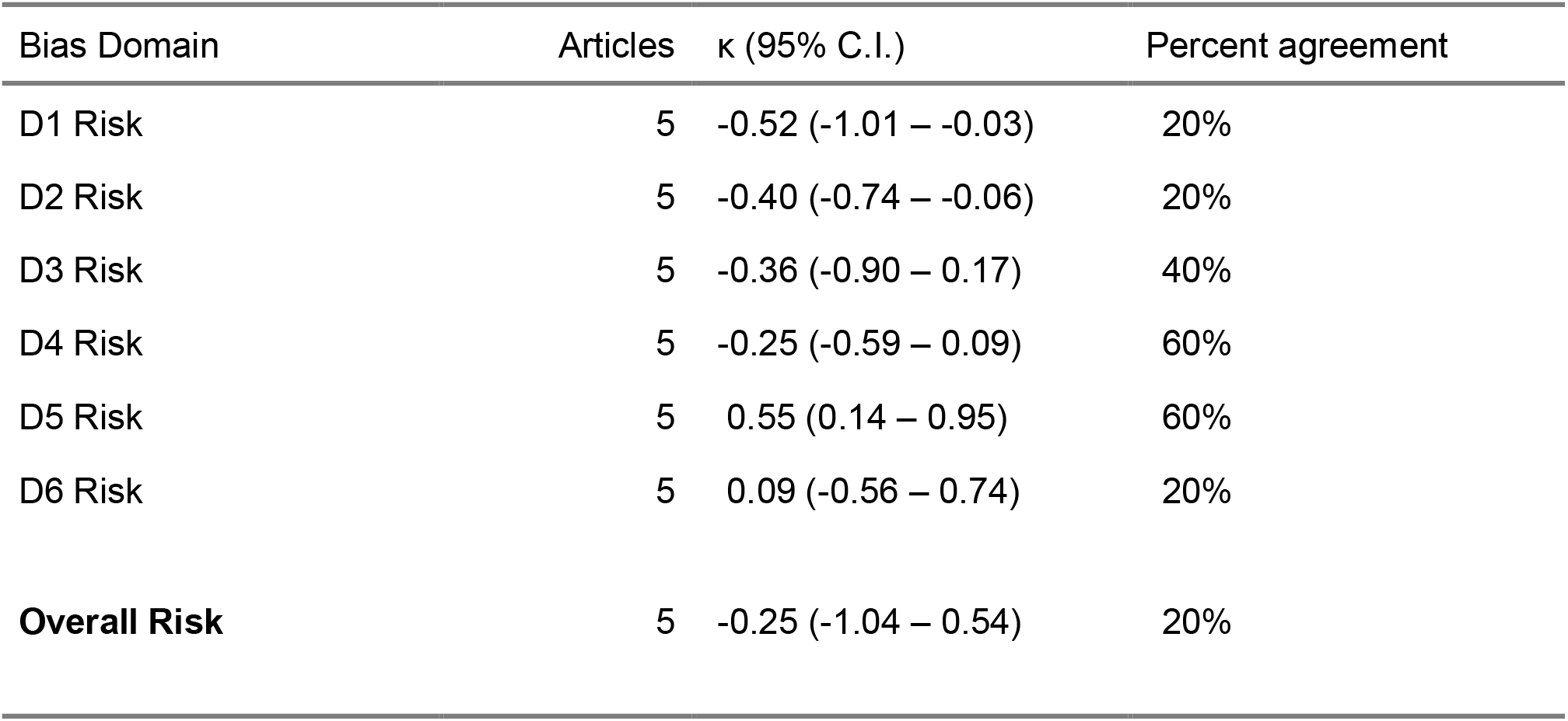
Overview of weighted Cohen’s κ scores across bias domains for human-human agreement of the duplicate articles across reviews, penalizing discrepancies between ratings in a quadratic manner.

| Bias Domain | Articles | $\kappa$ (95% C.I.) | Percent agreement |
| --- | --- | --- | --- |
| D1 Risk | 5 | -0.52 (-1.01 – -0.03) | 20% |
| D2 Risk | 5 | -0.40 (-0.74 – -0.06) | 20% |
| D3 Risk | 5 | -0.36 (-0.90 – 0.17) | 40% |
| D4 Risk | 5 | -0.25 (-0.59 – 0.09) | 60% |
| D5 Risk | 5 | 0.55 (0.14 – 0.95) | 60% |
| D6 Risk | 5 | 0.09 (-0.56 – 0.74) | 20% |
| **Overall Risk** | 5 | -0.25 (-1.04 – 0.54) | 20% |

## Discussion

This pilot study demonstrates the feasibility of adopting an LLM for ROB assessments using the QUIPS tool for systematic reviews of prognosis studies in neurology. With carefully designed prompts, the model was able to assign ratings across bias-domains and retrieve verbatim information to support manual verification.

### Limited human-LLM inter-rater agreement

Agreement for overall ROB was limited between human and LLM assessments (Cohen’s weighted κ, 0.22, 95% C.I.’s, 0.12–0.33). Cohen’s weighted κ showed considerable variation across both bias-domains and systematic reviews, ranging from 0.04 to 0.27 and from -0.15 to 0.45, respectively. Rank-based tests demonstrated statistically significant group-level differences across most domains, with LLM systematically assigning lower ROB scores.

To our knowledge, our present study with the QUIPS tool for neurological prognosis studies is the first of its kind, precluding comparisons with earlier findings. However, previous research on the agreement between human and LLM based ROB assessments - not using the QUIPS tool - shows heterogeneous results ^39,41,78–82^. Using a Claude 2 model, Eisele-Metzger found comparable agreement (Cohen’s κ of 0.22 for the overall score, and ranging from 0.10 to 0.31 across domains) ^41^. Similarly, a study found human-ChatGPT agreement using Cohen’s κ for overall ROB to range between 0.11 and 0.17, across various modelling strategies ^83^. Curiously, superior agreement was seen in a study by Lai et al., with Cohen’s κ ranging from 0.54 to 0.96 across multiple domains ^39^. As Eisele-Metzger et al. mention ^41^, these results may partly be explained by several considerations: 1) the authors used a modified ROB tool, which may be simpler to score; 2) only 30 articles were screened, and while randomly selected, this may lead to selection bias; and 3) the authors employed three methodological experts to independently rate ROB, rather than relying on previously published assessments.

To date, LLM for ROB assessment has mostly been explored in systematic reviews of randomized controlled studies (e.g., using ROB1 or ROB2 tools). Recently, this has been studied in two observational cohort studies ^84,85^, using the Newcastle-Ottawa Scale for observational studies ^86^. Results are inconsistent, with Xia et al. ^85^ reporting strong agreement and Beber et al. ^84^ mostly reporting poor agreement, though differing outcome measures complicate direct comparisons. Large methodological differences may account for these discrepant results: while Beber et al. used the original as-published ROB assessments, Xia et al. enlisted “three evidence-based medicine experts” to perform the ROB assessments to establish a “gold standard”. Other factors contributing to mixed findings likely include differences in prompt engineering (i.e., we used “zero-shot” prompting) ^36^, model type (e.g., “reasoning-enabled” versus “non-reasoning”) ^82^, and heterogeneous implementation of ROB frameworks (as seen in our study **S1**).

### Limited human-human inter-rater agreement

A noteworthy preliminary finding is that while LLM-human agreement was limited (κ = 0.22), the human-human agreement we could measure was even lower (κ = -0.25). However, the human-human subanalysis comprised only five article pairs, with a 95% CI spanning nearly the full theoretical range. Furthermore, the negative κ values for Domains 3 and 4, despite moderate percentage agreement, likely reflect the kappa paradox in small samples sizes with skewed categorical distributions. This comparison should therefore be considered hypothesis-generating rather than conclusive.

Furthermore, this finding substantiates previous doubts regarding treating published human ROB assessments as a definitive “gold standard” ^1,41,82^, with previous studies using ROB and ROB-2 tools demonstrating κ coefficients for overall risk score agreement as low as 0.02 ^27^ or 0.16 ^25^. Although earlier evidence on human inter-rater agreement for the QUIPS tool specifically showed higher scores - median κ score of 0.75 across nine review teams ^49^, and κ score for overall risk of bias of 0.48 ^50^ and 0.49 ^87^ – these assessments were done within the same team using the same protocol. By contrast, our human-human analyses compared articles rated across different research groups, with potentially different implementations (**S1**). Prior work from Hartling et al ^26^., Saadi et al ^88^., and Armijo-Olivo ^27^, albeit with different ROB tools, lends further evidence to the observation that ROB agreement is inferior across groups than within groups. Beyond concerns of an “imperfect reference standard” ^1^ for LLM validation, which affirms the need for validation studies using expert-derived ROB ratings, these findings suggest that ROB assessments in some published reviews may be unreliable.

## Sources of incongruency

Our inspection of “two-level” incongruencies for the Cochrane studies revealed no hallucinations and demonstrated that the LLM was able to provide thorough and reliable supporting statements. In three out of four cases, we agreed with the LLM’s overall ROB assessment. This contrasts with Eisele-Metzger et al.^41^, who never agreed with LLM ratings (Claude) when inspecting “two-level” discrepancies. Possible explanations for this contrast include: 1) our scoring algorithm heavily penalized “High” ratings (i.e., even when only one domain is rated as “High”, then the overall score automatically becomes “High”) and 2) the observation that our LLM-pipeline systematically assigned lower ROB assessments than published human assessments. For Adan et al.^77^, for instance, the Cochrane reviews consistently considered the study population (D1) to have a higher ROB than the LLM. This conflicts with earlier work that found LLMs to instead have a “. . . tendency to overclassify ROB . . . ” ^82^.

Another notable finding was that the human reviewers from West et al. ^53^ consistently assigned higher ROB ratings for Domain 6 than the LLM, attributing a “High” ROB to 12/24 articles (vs. 0/24 articles). In contrast to the LLM, humans commonly assigned a “High” score for analyses where “. . . only significant variables entered into the model”. Although this conventional analytic approach admittedly has shortcomings, in our view this should not automatically warrant a “High” risk of bias. This and other discrepancies may underlie the exceptionally poor agreement (κ 0.03) seen in this domain. Van Grooten et al., similarly found inter-rater agreement in this domain to be lower compared to most other domains, possibly because this domain relies strongly on methodological expertise and personal preferences ^50^. Because West et al. did not contribute to inter-rater agreement calculations for the overall ROB assessment (except for one article) due to missing ratings from certain domains - in addition to Giuliano et al., - overall inter-rater agreement between human and LLM assessments may therefore be artificially inflated.

## Strengths and limitations

A key strength of this study is that our dataset and LLM-pipeline are open-source and freely accessible, facilitating replication and future experimentation. Furthermore, our consistent deterministic scoring algorithm and study design enabled head-to-head comparisons between human and LLM assigned ROB assessments, and between human pairs.

An important limitation is our small sample size — particularly the human-human subanalysis (n = 5 article pairs) — despite the overall corpus (298 articles) being substantially larger than similar studies ^39,41^. Larger scale studies are needed to enable identification of sources and areas of agreement and disagreement between human and LLM ratings, as well as between human pairs. In addition, as human-human inter-rater agreement may decline with lower-quality studies ^50^, future work should investigate whether human-LLM agreement is similarly influenced by the overall quality of studies or systematic reviews in general (for example, in high-quality Cochrane Systematic Reviews). Alas, our limited sample size did not allow us to study this, and hence no conclusions can be made regarding the considerable variation in inter-rater agreement across studies, such as between McGeown (κ 0.44) and Thiara (κ -0.15). Additionally, a challenge inherent to this area of research is that both misunderstanding of 1) ROB criteria or 2) article content may contribute to inter-rater variability, and separating their relative contributions is infeasible ^50^.

Another limitation is that systematic reviews differed in their application of the QUIPS tool (see ***S1B***); although in the original QUIPS documentation modifications are encouraged when appropriate ^49^, they complicate comparisons in this study. Some studies aggregated scores in accordance with earlier recommendations ^50^, while others implemented custom aggregation approaches, and still others did not aggregate scores at all. In addition, some systematic reviews substituted or excluded domains entirely ^52,77^. To illustrate, Adan et al., replaced “Study Confounding” with “Adjustment for Covariates”, which though reasonable, may introduce inconsistencies as it remains unknown how alternative QUIPS approaches affect inter-rater agreement ^50^. Furthermore, West et al., and Giuliano et al., did not rate certain domains, and hence no aggregated overall risk scores were possible; conversely, Adan et al. rated this category as “Unclear”, which we interpreted as “Moderate”, enabling aggregation. Finally, due to absence of supporting documentation for non-Cochrane systematic reviews in this study, we were unable to compare the justifications for ratings provided by the original human ratings with those of the LLM in 52/56 cases. This hinders investigations into incongruencies, and in the formal QUIPS documentation Hayden et al. advised supporting judgements with information verbatim from the text^49^.

Overall, our findings signal the need for updated QUIPS documentation, where potential modifications/deviations are standardized, in addition to detailed guidance for deriving aggregate scores ^24,50^. This issue gains urgency as LLMs become increasingly available for assistance in ROB assessment ^36^.

## Future directions and potential applications

The volume of systematic reviews continues to explode ^89,90^. Considering the rapid adoption and development of automated tools for all stages of systematic reviews ^91^, we reiterate the need for ^1,41,83^ broad dialogue to standardize methods for validating and reporting LLM use in ROB assessments, including which outcome measures to report ^29,36,37,85^. Crucially, future research, perhaps in the form of an umbrella review, should study the causes and areas of heterogeneous results within-studies (e.g., across bias domains) and between-studies ^1^. As LLMs continue to evolve, we concur with Sahu’s recommendations that frameworks should be developed for monitoring performance between models and over time, and that authors should explicitly describe which models and versions were used to boost reproducibility ^1^.

Recognizing that differences in prompt design can lead to variable results ^36,92^, this too should be standardized and reported transparently. Methods should be developed to reliably quantify and compare time and costs saved across different tasks within systematic reviews ^48^, in order to determine their relative benefit. For clinical research in neurology, prospective efforts should seek to identify challenges and objections to the routine implementation of LLMs in systematic research workflows ^28,93^.

With refinement and validation, our LLM-based tool has the potential to become a valuable tool for clinical researchers conducting systematic reviews in the field of neurology^94^. Initially the focus should be on how automated technology can assist human reviewers, rather than replace them completely ^1,28,40,82,93^. Nyrhi et al. proposed two possible roles for LLMs in ROB assessment: 1) as a screening/flagging instrument for “High” risk articles; and 2) as “. . . the second assessor in a dual-review workflow”, with conflicts adjudicated by a third human (independent) assessor^82^. Another potential application of our tool could be automatically performing ROB assessments on a weekly or monthly basis for newly published nonrandomized prognosis articles, allowing for seamless incorporation into subsequent systematic reviews for neurology and beyond.

## Implications for Research – Cost and Time Savings

Automated tools for ROB assessment have the potential to conserve significant costs and time ^95^, and may enable the reallocation of resources to more complex or collaborative tasks ^29^. A hypothetical calculation follows, using a previous estimate that ROB assessment using QUIPS has a median completion time of 20 minutes ^45^. For a systematic review with 20 studies and two independent assessors ^50^, this translates to ∼ 13 hours of labor. Given an estimated hourly rate (e.g. PhD-student or resident) of $50 USD, this amounts to ∼$650 USD per systematic review. A brief search on PubMed (April 13, 2026; ("traumatic brain injury"[Title/Abstract] OR "epilepsy"[Title/Abstract] OR "stroke"[Title/Abstract] OR "Parkinson"[Title/Abstract] OR "dementia"[Title/Abstract] OR "neuropathy"[Title/Abstract] OR "migraine"[Title/Abstract] OR "neurolog*"[Title/Abstract]) AND ("systematic review*"[Title/Abstract] OR "meta-analysis"[Title/Abstract]) AND ("prognosis"[Title/Abstract] OR "prognostic"[Title/Abstract])) finds ∼430 articles published in 2025, suggesting that in one year ∼$300,000 USD in *funding may have indirectly been allocated towards this task for neurological prognosis systematic reviews alone.* This is a conservative estimate, as it excludes training, conflict resolution, or involvement of senior clinicians or researchers with higher hourly rates. While total replacement of human reviewers with an automated tool requires further validation and evidence of acceptable inter-rater agreement, this estimate serves to illustrate the scale of this issue.

## Conclusions

We present findings of a pilot study using a tailored LLM applying the QUIPS framework for ROB assessments in neurology prognostic systematic reviews. Although agreement between human- and LLM-derived ROB assessments was limited, our findings suggest that this performance is not inferior to ratings between human pairs. While refinements are needed, the integration of automated ROB assessments in systematic reviews is an exciting prospect, including for prognosis studies in neurology and beyond.

## Data availability

Our data, code, and LLM-based pipeline are available in a GitHub repository: https://github.com/SemKampman/QUIPS_Automated_ROB_Assessment.

## CRediT

SK: Conceptualization, methodology, data collection, formal analysis, software and writing – original draft.

KB: Conceptualization, supervision, and writing – revision.

WO: Conceptualization, supervision, methodology, formal analysis, software and writing – revision.

## A.I. usage declaration

Beyond the use of LLMs as described in the Methods, A.I. tools were used throughout manuscript preparation to improve readability, coherence and overall flow. In addition, the Paper Assessment Tool (https://github.com/bdsp-core/PAT-PaperAssessmentTool) was used to proofread the manuscript and identify areas for improvement prior to submission. A.I. tools were also used to generate and debug R code for statistical analyses. Claude Sonnet 5 (*Anthropic*) and Microsoft 365 Copilot (Microsoft Corporation) were used for these purposes. Authors reviewed and verified all generated content.

