## Supplemental Material for "A large language model for risk-of-bias assessment in systematic reviews of prognosis studies in clinical neurology"

**S1A.** An overview of included systematic reviews, with key objectives and number of included studies listed. TBI = traumatic brain injury. The key objectives were adapted from the original studies, with some text retained verbatim and some partly summarized.

### A. Study Characteristics

| Study ID | Journal of publication | Key objectives | No. of included studies |
|---|---|---|---|
| | | **Epilepsy (n = 94)** | |
| **Adan 2025** | *Cochrane Database of Systematic Reviews* | To assess prognostic factors associated with seizure recurrence/epilepsy diagnosis following either a first unprovoked seizure, a seizure cluster, or first presentation of status epilepticus. | 23 |
| **Arfaie 2023** | *Seizure: European Journal of Epilepsy* | To assess pre-operative and post-operative intelligence outcomes in children subject to epilepsy surgery. | 7 |
| **Giuliano 2021** | *Seizure: European Journal of Epilepsy* | To assess differences in prognostic factors and long-term outcomes between male and female patients in juvenile myoclonic epilepsy. | 25 |
| **West 2019** | *Cochrane Database of Systematic Reviews* | To assess epilepsy surgery outcome and factors associated with post-operative seizure remission. | 182 (of which only 28 were assessed with the QUIPS tool) |
| **Wassenaar 2013** | *Epilepsy Research* | To assess prognostic factors for intractability in epilepsy. | 11 |
| | | **Stroke (n = 79)** | |
| **Gallucci 2026** | *International Journal of Stroke* | To assess the association between cognitive reserve and stroke outcome | 17 |
| **Pires 2025** | *Neuroradiology* | To assess the association between diffusion kurtosis imaging markers and stroke outcomes | 11 |

| | | | |
|---|---|---|---|
| **Zhubi 2025** | *Journal of Clinical Medicine* | To assess the association between FLAIR positivity and outcomes following intraveneous thrombolysis in known-onset strokes. | 6 |
| **Thiara 2024** | *Critical Care Medicine* | To assess predictors of intracranial hemorrhage and ischemic stroke in adult patients subject to venovenous extracorporeal membrane oxygenation. | 17 |
| **Tao 2023** | *Frontiers in Neurology* | To assess the impact of cognitive reserve sociobehavioral proxies on ischemic stroke outcomes. | 28 |
| | | **TBI (n=125)** | |
| **Verboom 2026** | *Journal of Neurotrauma* | To assess the prognostic value of EEG markers in the acute phase in TBI patients in the ICU. | 27 (of which 23 were assessed with the QUIPS tool; only 22 were available for analysis, as Beridze et al., 2010, was not amenable to markdown conversion) |
| **Dery 2025** | *Neurotrauma reports* | To assess the prevalence of symptom resolution in non-athletic TBIs, and to assess factors associated with recovery. | 16 |
| **McGeown 2025** | *NeuroImage: Clinical* | To assess associations between MRI features and symptom burden of mild TBI and subsequent functional recovery. | 62 (of which only 61 were available for analysis; one article was embargoed [Onicas et al., 2025]) |
| **Tan 2025** | *European Geriatric Medicine* | To assess the functional recovery in older adults after a mild TBI. | 18 |
| **Ghozy 2022** | *Frontiers in Neurology* | To assess the effectiveness of the neutrophil-to-lymphocyte ratio for predicting TBI outcomes. | 8 |

**S1B.** An overview of QUIPS implementation approaches. QUIPS assessment, modification and methods for deriving overall score were adapted from the original text, with some sections reproduced verbatim and others summarized.

### B. QUIPS implementation

**Epilepsy systematic reviews**

| Study ID | QUIPS assessment | QUIPS modifications and other notes | Method for deriving overall score |
|---|---|---|---|
| Adan 2025 | Independently evaluated by two authors. | "Confounding'" domain replaced with "adjustment for other prognostic Factors". Further, the authors use "Unclear", either instead of "Moderate" or "N/A". *****<br><br>Though for overall risk of bias per article, they seem to equate "Unclear" with "Moderate". | "We considered studies to have an overall low risk of bias if most QUIPS domains (i.e. at least four of six) were rated as low risk and none were rated as high risk" |
| Arfaie 2023 | Evaluated by two reviewers, and verified by a third reviewer. | Not reported. | "Mean risk scores for each domain were calculated by associating the level of risk with numbers (low = 1, moderate = 2, high = 3)." |
| Giuliano 2021 | Independently evaluated by two authors. | "The study attrition subscale was not used as it was not always applicable" | "If all the domains were classified as having a low RoB or included just one with a moderate RoB, then the article was defined 'low RoB'; if one or more domains were classified as having a high RoB or >3 as having a moderate RoB, then the article was defined 'high RoB'. All papers falling between these criteria were defined 'moderate RoB'." ***** |
| West 2019 | Two review authors assessed risk of bias, and two review authors independently checked these judgements. | Rated Domain 2 (Study Attrition) as N/A for retrospective/registry studies. Rated Domain 5 (Study Confounding) as N/A ". . . for studies of a single-group design", | Not performed. |
| Wassenaar 2013 | Not specified. | Not reported . | Not performed. |

**Stroke systematic reviews**

| Study ID | QUIPS assessment | QUIPS modifications and other notes | Method for deriving overall score |
|---|---|---|---|
| Gallucci 2026 | Independently evaluated by two authors. | "Studies with a high risk of bias were excluded from the meta-analysis. We further excluded two studies with only moderate bias, due to a reported strong association between | Though aggregation was done, the approach was not explicitly described. However, it can be deduced that more lenient methods were used than in Grooten et al. For example, articles with **≥2** domains rated as "Moderate" were rated overall as being |

| | | | |
|---|---|---|---|
| | | CR-proxy (lower level of education) and restricted health care access in these studies, providing the unresolved bias on functional outcome” | “Low”, and articles with more >3 domains as “Moderate” or at least one domain of “High” were still given an overall rating of “Moderate”. |
| Pires 2025 | Independently evaluated by two authors. | Not reported. | “Based on all six domain results, the overall RoB of the study was derived.”<br><br>However, it is unclear how the authors derived the total score. It appears more lenient than recommended, as for example one high domain did not automatically result in a high overall risk score. |
| Zhubi 2025 | Independently evaluated by two authors. | Not reported. | Method of aggregation not mentioned in the text. However, it can be deduced from supplementary that a very strict approach was used, though not corresponding to Grooten et al.’s approach. For instance, an article with one domain rates as moderate is here given an overall rating of moderate. |
| Tao 2023 | Not specified. | Not reported. | Though aggregation was done, the approach was not explicitly described. However, it can be deduced that it was more lenient than recommended by Grooten et al. For example, an article with three domains rates as moderate was given an overall estimate of moderate. |
| Thiara 2024 | Evaluated by two reviewers. Unclear if this was done in an independent or blinded fashion. | Not reported. | “Consistent with QUIPS guidelines, observational studies start as having a high risk of bias based on study design, and can be downgraded . . . based on factors …”<br><br>To our knowledge, however, this is not consistent with QUIPS guidelines. However, despite specifying this in the methods, no efforts are actually made to aggregate these scores in the manuscript. The authors subsequently use risk-of-bias to inform GRADE certainty assessment, grading some articles as “Serious” and others as “Not serious”, though it is unclear how this score was derived and how this influenced certainty assessment. |

**TBI systematic reviews**

| Study ID | QUIPS assessment | QUIPS modifications and other notes | Method for deriving overall score |
|---|---|---|---|
| Verboom 2026 | Not specified. | Not reported. | Not performed. |
| Dery 2025 | Evaluated by two reviewers. Unclear if this was done in an independent or blinded fashion. | Not reported. | “Following the assessment of the six domains, a judgement was made om the overall risk of bias of the study using the same ratings.” However, the exact approach is not clearly articulated. Further, the approach seems more lenient than Grooten et al., recommendations, with two domains rated as High still receiving overall rating of “Moderate”. Further, there seem to be internal inconsistencies. For instance, while most studies with two “Moderate” and one “High” risk ratings, receive an overall rating of “Moderate”, one received an overall rating of “High”. |

| | | | |
|---|---|---|---|
| McGeown 2025 | One author evaluated all articles, with a second author randomly verifying 10%. | Not reported. | No aggregation done. In fact, the authors recognize Hayden et al.s' original guidance: "Consistent with guidance from the developers, no single summary score is presented for each study". |
| Ghozy 2025 | Independently evaluated by three authors. | Rather than using "Low" , "Moderate" or "High" risk-of-bias, the authors use terms such as "Yes", "No", "Partly" or "Unsure". | Not performed. |
| Tan | Independently evaluated by two authors. | Not reported. | Though aggregation was done, the approach was not explicitly described. However, it can be deduced that it was more lenient than recommended by Grooten et al. For example, an article with > three domains rated as "Moderate" or one domain rated as "High" still received an overall rating of "Moderate". |

**S2.** Study investigators (SK, WO) attempted to reconcile differences between human and LLM ratings for overall risk-of-bias scores. We studied articles with “two-level” overall risk-of-bias differences (e.g., “High” vs “Low”) included in Cochrane systematic reviews because these provide supporting statements for risk-of-bias assessment.

| Review | Article | Elaboration Cochrane | Elaboration LLM | Author’s judgement (SK and WO, unblinded) |
|---|---|---|---|---|
| Adan 2025 | Hauser 1990 | D1 = High; “Very narrow inclusion criteria - recruitment had to be within 24 hours”<br>D2 = Low; “20 participants out of 208 withdrew from the study”<br>D3 = Low; “Well described”<br>D4 = Low; “Well represented”<br>D5* = Unclear †; “Some attempt to adjust made”<br>D6 = Low; “Appropriate methods used”<br><br>Overall, Cochrane = Unclear/Moderate ✚<br>Overall, using our algorithm = High<br><br>* : Adan et al. replaced “Study confounding” with “Adjustment for other prognostic factors”<br>†/✚ : Adan et al., used “Unclear” when “Low” or “High” was not applicable. We interpreted this as “Moderate”. | D1 = Low; “The study used a robust surveillance system to identify patients at the point of first medical contact, minimizing referral bias. Inclusion/exclusion criteria were highly specific and the baseline characteristics were well-defined.”<br>D2 = Low; “The attrition rate was relatively low (approx 15%), and the authors noted that most dropouts contributed significant follow-up time. Reasons for withdrawal were documented.”<br>D3 = Low; “Prognostic factors were defined using international standards (ILAE) and measured at baseline, preventing knowledge of the outcome from influencing the measurement.“<br>D4 = Moderate; “While the outcome (seizure recurrence) is clearly defined, it relies on patient/parent report during 6-12 month intervals, which may be subject to recall bias, and there is no mention of blinding the outcome assessors to the baseline PFs.”<br>D5* = Low; “The study used multivariable Cox proportional hazards models to adjust for potential confounders, including etiology and EEG findings.”<br>D6 = Low; “The statistical methods (Life-table, Kaplan-Meier, Cox regression) are appropriate for time-to-event data. The reporting is transparent, including both univariate and multivariate results.”<br><br>Overall = Low | D1 = “Moderate”; We don’t consider the narrow inclusion criteria to substantially increase the risk of bias for this domain, although this may have jeopardized the representativeness of the study sample for the population of interest.<br>D2 = “Low”; The loss-to-follow up and withdrawal burden seems acceptable and is adequately explained. As noted by the LLM, most patients who were lost to follow up still contributed to the 2-year outcome.<br>D3 = “Low”; We agree with both human and LLM assessments.<br>D4 =”Low”; We agree with the human assessment. The outcome is well-defined, easy to measure, consistently applied across participants. Given the regularity with which patients were contacted and the salience of paroxysmal events, we consider the potential impact of recall bias to be minimal.<br>D5 = “Low”: We agree with the LLM assessment. Furthermore, it is unclear what exactly the human assessors are concerned about.<br>D6 = “Low”; We agree with both the human and LLM assessments.<br><br>Overall = Low |

| | | | | |
|---|---|---|---|---|
| Adan 2025 | Mahler 2015 | D1 = High; “Clearly defined retrospective study, some conflation of single seizure and multiple seizure presentations”<br>D2 = Unclear; “N/A - retrospective registry study”<br>D3 = High; “Mostly focussed on a single prognostic factor - TBI, only some mention of other factors”<br>D4 = Unclear; “Appropriately defined outcomes and end points for the study but within a very narrow spectrum of TBI”<br>D5 = Unclear*; “Some sensitivity checks for possible confounding factors”<br>D6 = Unclear; “Appropriate statistical reporting with risk ratios stated clearly from univariable statistics but no multivariable model used in the paper”<br><br>Overall, Cochrane = High<br>Overall, using our algorithm = High | D1 = Low; “ The study uses a robust population-based registry (SIRE) and the Swedish National Inpatient Registry, covering a well-defined urban population of nearly 1 million. Recruitment was comprehensive, involving multiple reporting mechanisms and central EEG screening, ensuring the sample is highly representative of the target population.”<br>D2 = Low; “As a registry-based case-control study using mandatory national registries (IPR), the risk of attrition is minimal. The study captures all hospitalized TBI events and incident seizures within the registry period without the traditional 'loss to follow-up' seen in prospective cohorts.”<br>D3 = Moderate; “While TBI is defined by ICD codes in a mandatory registry, the authors acknowledge that the severity categorization is 'crude and arbitrary' and relies on hospitalization. This likely misses non-hospitalized mild TBIs, potentially biasing the risk estimate for the 'mild' category.”<br>D4 = Low; “The outcome (unprovoked seizure) was rigorously validated by a multi-disciplinary panel of experts reviewing medical records, which is the gold standard for epilepsy incidence studies.”<br>D5* = Low; “The study matched controls by age, sex, and location. Furthermore, it performed a sensitivity analysis adjusting for major confounders (alcohol, stroke, dementia, psychiatric disease) and found they did not significantly alter the results.“<br>D6 = Low; “The study uses appropriate conditional logistic regression for the matched case-control design. It provides detailed subgroup analyses (time since injury, TBI type) and reports confidence intervals for all major findings. There is no evidence of selective reporting.”<br><br>Overall = Low | D1 = Moderate; Due to limited descriptions there is indeed a possibility that single seizure and multiple seizure presentations are conflated in this study. However, given the study objective, this does not per se mean that the sample population is not generalizable. Furthermore, the Cochrane authors could question whether this study fulfills their own PICO-framework and inclusion criteria. Perhaps the authors are conflating their own uncertainty over whether the article is eligible with the question of whether the study sample reflects the greater population of interest.<br>D2 = Moderate; The scoring of this domain for retrospective/registry studies represents a quandary as no clear guidelines exist for how to rate study attrition for this study design. However, using Hayden et al.’s criteria, we do not consider this to constitute either a high or low risk of bias, but do acknowledge that there may be differences in the participants who do and do not have adequate follow-up.<br>D3 = High: We agree with both the reasoning of the human and LLM assessors, but consider a high risk of bias to be present.<br>D4 = Low; We agree with the LLM assessment. The fact that this outcome is studied within only a narrow spectrum of TBI is, in our view, not a reason to avoid giving a Low score for this domain.<br>D5 = Low; We agree with the LLM assessment as the authors attempted to minimize bias in this domain despite the case-control study design.<br>D6 = Moderate; We agree with the human assessors.<br><br>Overall = High |
| Adan 2025 | Takami 2021 | D1 = High; “Only children who had an EEG were included in the study”<br>D2 = Unclear; “No mention of study attrition or dropouts from study”<br>D3 = Low; “High number of prognostic factors collected as part of study” | D1 = Low; “The study provides a clear description of the target population, recruitment setting, and inclusion/exclusion criteria. The sample size is large for a single-center study, and the recruitment process appears systematic.”<br>D2 = Low; “The study reports a very high follow-up rate for a long-term study, with over 95% of patients followed | D1 = Low; It is very typical to receive an EEG after a first seizure as it forms an integral component of the diagnostic workup.<br>D2 = Moderate; While the authors describe sound methods for ascertaining follow-up information and report high follow-up rates, no reasons or timelines for dropouts are given. |

| | | | | |
|---|---|---|---|---|
| | | D4 = Low; "Clear study end points and outcomes clearly defined"<br>D5 = Unclear; "No formal reporting of study confounders but they do use multivariable models to control for a number of variables"<br>D6 = Low; "Appropriate use of univariable and multivariable statistics"<br><br>Overall, Cochrane = Unclear/moderate<br>Overall, using our algorithm = High | for at least 2 years. The methods for follow-up (visits or telephone) are appropriate."<br>D3 = Low; " Prognostic factors (EEG, neuroimaging, etiology) were measured using standard clinical tools and reviewed by specialists. The definitions used are consistent with ILAE guidelines."<br>D4 = Moderate; "The outcome (recurrence) is clearly defined, but its measurement relies partly on parental recall via telephone interviews, which introduces a risk of recall bias, although this is standard in epilepsy prognosis studies."<br>D5* = Low; "The study used multivariable Cox proportional hazards modeling to adjust for a wide range of potential confounders, including age, sex, seizure type, and etiology."<br>D6 = Low; "The statistical methods (Kaplan-Meier and Cox regression) are standard and appropriate for time-to-event data. The results are reported transparently with confidence intervals."<br><br>Overall = Low | D3 = Low; We agree with both human and LLM assessments.<br>D4 = Low; We agree with the human assessment. We consider the risk of recall bias to be minimally present.<br>D5 = Low; Although we agree with the human assessors that formal reporting of study confounders would have been preferred, their multivariable modelling approach adjusts for many potential confounders and is adequately reported.<br>D6 = Low; We agree with both human and LLM assessments.<br><br>Overall = Low |
| West 2019 | Schramm 2011<br><br>**Note**; given this was an interventional RCT, the Cochrane Risk of Bias tool (ROB 2) would have been more appropriate. | D1 = Moderate; "Large sample recruited; seems representative of the population of interest; however 35% of potentially eligible individuals not included for varying reasons"<br>D2 = Low; "All participants in the study contributed data from 1 year and were included in an intention-to – treat analysis; secondary analysis was planned in the case of dropouts"<br>D3 = Moderate; "Clinical/demographic data seem to have been collected in a reliable and valid way under the same protocol for all patients; however unclear what are prognostic variables and what are confounding factors""<br>D4 = Low; "Difficult outcome to measure in a reliable and valid way but seems satisfactory; measured by a known scale at a single time point in the same way for all participants<br>D5 = Moderate; "Clinical/demographic data seem to have been collected in a reliable and valid way under the same protocol for all participants; however unclear what are prognostic variables and what are confounding factors; "confounders" | D1 = Low; "The study provides a clear description of the target population, recruitment centers, and inclusion/exclusion criteria. A detailed flowchart accounts for eligible non-participants, and baseline characteristics are well-balanced between groups.<br>D2 = Low; "The study achieved 100% follow-up for the primary outcome (seizure freedom at 1 year) for all 207 randomized patients, using an intent-to-treat approach."<br>D3 = Low; "The prognostic factor (intended resection length) was the intervention itself in this RCT. It was clearly defined and verified postoperatively using objective MRI volumetry with high inter-rater reliability."<br>D4 = Low; "The outcome (seizure freedom) was measured using the standard Engel scale. Assessors were blinded to the resection group, and follow-up intervals were standardized."<br>D5* = Low; "As an RCT, randomization successfully balanced known confounders (age, duration, pathology). Multivariable analysis was not the primary focus, but the balance of baseline characteristics suggests low risk." | D1 = Low; We agree with the LLM assessment. Although a high % of patients are excluded, these reasons are well-documented and understandable, and do not invalidate the sample population's representativeness of the population of interest.<br>D2 = Low; We agree with both human and LLM assessments.<br>D3 = Low; We agree with the LLM assessment.<br>D4 = Low; We agree with both LLM and human assessors.<br>D5 = Low; We agree with the LLM assessment. In a related study ("Part 2: volumetric resection extent and subgroup analyses") the effects of confounding variables are explored. Interestingly, the humans do not refer to this, though they would have had access to these materials, while the LLM did not.<br>D6 = Low; Again, we agree with the LLM assessment.<br><br>Overall = Low |

| | | | |
|---|---|---|---|
| | | do not appear to be accounted for in the design, and unclear what type of analysis (if any) was done"<br>D6 = Moderate; "Unclear which variables have been included in the model, exactly what type of analysis has been performed, and whether all results have been reported"<br><br>Overall, Cochrane = did not aggregate<br>Overall, using our algorithm = High | D6 = Low; "The statistical plan is well-described, including power calculations and an ITT approach. Results are reported transparently for all randomized patients."<br><br>Overall = Low |
